# Filum Terminale Diameter on Routine Pediatric MRI: A Large-Cohort Clinical Reference in 3,406 Children and the Age-Dependent Meaning of the 2-mm Thickened-Filum Threshold

**DOI:** 10.64898/2026.06.14.26355614

**Authors:** Wenwen Tang, Yuan Dong, Jinliang Chen, Yue Yang, Hong Huang, Mengyan Yu, Jun Zhu, Gang Shen

## Abstract

**Background:** A filum diameter >2 mm is the conventional MRI threshold for a thickened filum, but it derives from small, mostly adult series showing no age dependence; whether one cutoff suits all of childhood is untested.

**Objective:** To build an age-specific filum-diameter reference on routine pediatric MRI and test, adjusting for image resolution, whether the 2-mm threshold is age-stationary.

**Materials and methods:** In this retrospective study an nnU-Net tracer measured the maximal filum diameter on consecutive lumbosacral MRI; versus manual tracing it showed negligible bias but moderate single-measure agreement. After excluding report-confirmed fatty filum, lipoma, or tethered cord, the proportion >2 mm was analysed within one acquisition protocol and by logistic regression adjusting for voxel size and slice thickness.

**Results:** Of 7,245 examinations, 3,869 (53%) were traceable; untraced ones were younger (median 0.75 vs 2.0 years). The presumed-normal cohort had median diameter 1.48 mm. At matched resolution, 2 mm marked the 94th percentile in infants (5.6% exceeded it) but the 83rd by 3–6 years (17.4%); the age effect persisted after adjusting for voxel size and slice thickness (3–6 years vs infants, adjusted OR 4.7; P < .001).

**Conclusion:** Filum diameter clusters near 1.5 mm, and the fixed 2-mm cutoff flags ∼5% of infants but ∼17% of preschoolers. Caliber should be judged against an age-specific clinical reference, not one fixed cutoff; a thick filum is not itself a diagnosis of tethered cord.

## Introduction

The filum terminale is a thin fibrous strand extending from the conus medullaris to the dural sac; its abnormal thickening or fatty infiltration is a recognized substrate of the tethered cord spectrum and a target for surgical sectioning [1–3]. On MRI a filum is conventionally called “thickened” when its diameter exceeds 2 mm. The morphometric basis for this cutoff rests on small series: in the largest MRI plus cadaveric study the mean filum diameter was ∼1.5–1.6 mm, a diameter >2 mm occurred in only 7–8%, and — importantly — caliber showed no age or sex dependence in adults [4]. Whether the same fixed 2-mm boundary applies to a one-year-old and a teenager has not been tested against a large pediatric reference.

Two gaps motivate this work. First, large-scale pediatric reference data for filum diameter are scarce; most measurements rest on tens to low hundreds of examinations [4]. Our companion conus-position reference established a population-scale distribution and showed that many surgically treated children have a normally positioned conus [6], redirecting attention to the filum and to quantitative signs that may be tested separately. A conus-radiomics companion manuscript has been submitted separately, and two additional registry analyses (conus longitudinal reproducibility; filum T2-signal) remain unpublished; all are available from the authors on request. The filum-diameter reference and age-dependent 2-mm threshold analysis reported here have a distinct endpoint and do not duplicate the primary results, tables, or figures of those companion analyses. Second, the filum is small and is seldom quantified in routine reports, so its caliber is rarely measured in practice. An automated, objective measurement deployed at cohort scale can address both gaps — provided the measure is validated and the analysis controls for the image-resolution effects that dominate a structure only a few voxels wide.

We therefore deployed a validated automated filum tracer across a large pediatric lumbosacral MRI registry to (a) establish an age-specific reference distribution for filum diameter, (b) test, with explicit adjustment for image resolution, whether the fixed 2-mm threshold has a stationary percentile meaning across childhood, and (c) compare objective measurement against free-text reporting of filum thickening.

## Materials and Methods

Study design and ethics. This retrospective single-center study was approved by the Ethics Committee of Women and Children’s Hospital of Ningbo University (approval no. NBFE-2026-KY-130; expedited review; approved 2 June 2026), which waived informed consent. Consecutive children (<16 years) who underwent lumbosacral MRI between January 2021 and May 2026 were eligible. The present analysis was conducted as a distinct filum-diameter reference and threshold study within the registry and does not share primary endpoints, tables, or figures with related registry analyses.

Imaging and automated filum tracing. The filum was assessed on the axial T2-weighted series (single-shot turbo spin-echo; in-plane voxel 0.5–0.66 mm; slice thickness median 6.5 mm, the routine clinical protocol). A 2-D nnU-Net [7] was trained to trace the filum cross-section on 76 manually annotated axial cases (drawn by an operator experienced in spinal MRI; a formal second-reader inter-rater study was not performed — see Limitations) and deployed across the registry so all examinations were measured identically. To check that the automated diameter reflects anatomy rather than a learned label, it was compared with manual tracing on the paired cases (Bland–Altman bias and limits of agreement; two-way random single-measure intraclass correlation, ICC). As a leakage sensitivity analysis, the reference percentiles were recomputed after excluding the tracer’s training cases.

Diameter measurement. On each trace the filum cross-sectional area was computed slice by slice from the in-plane voxel area, and the diameter was defined as the maximal single-slice equivalent diameter, 2×√(area/π) — the thickest cross-section, matching the convention a radiologist applies. Because the filum spans only 1–3 in-plane voxels, the diameter is quantized at approximately the in-plane voxel size (0.5–1.0 mm); we therefore report percentiles empirically and, critically, analyse the 2-mm threshold with explicit control for image resolution (below) rather than relying on a smooth model.

Reference cohort and groups. Examinations with a report-confirmed fatty filum, intradural/filar lipoma, or tethered cord were excluded to define a presumed-normal cohort; one examination per child (the earliest) was retained. Children with a report-confirmed fatty filum/lipoma, and those with a report of tethered cord, were analysed separately for comparison.

Resolution-controlled threshold analysis. Because in-plane voxel size both quantizes the diameter and varied modestly with age, the proportion exceeding 2 mm was assessed two ways: (i) within the single dominant acquisition protocol (in-plane voxel 0.64–0.67 mm, 81% of the cohort), removing between-protocol resolution differences; and (ii) by multivariable logistic regression of (diameter >2 mm) on age band with both in-plane voxel size and slice thickness as covariates. To confirm the age effect is not an artifact of the specific cutoff or of quantization in the borderline range, it was re-tested at 1.8, 2.0 and 2.2 mm and after excluding the 1.8–2.2-mm gray zone. We report age-band percentiles (median, 90th/95th/97th), the proportion >2 mm (Wilson 95% CI), the percentile rank of 2 mm, group differences (Mann–Whitney U), and a traced-versus-untraced comparison (Supplementary Table S1). Analyses used Python (NumPy, SciPy, statsmodels); P < .05 was considered significant.

## Results

Cohort and tracer validation. The tracer produced a valid filum trace in 3,869 of 7,245 deployed examinations (53%; the remainder had no resolvable axial filum, most often because the axial stack acquired for the conus did not extend to the lower filum). Against manual tracing the automated diameter showed negligible bias (−0.03 mm) but only moderate single-measure agreement (ICC 0.69; limits of agreement −0.53 to +0.48 mm, ≈ one voxel — large relative to the 2-mm threshold; n = 57 paired cases enriched for thick/fatty fila). The measure is therefore unbiased at the group level but imprecise per filum, which is why we anchor inference to age-band proportions and resolution-adjusted models rather than to individual diameters. Excluding the tracer’s training cases left the reference percentiles essentially unchanged (e.g., infant median 1.48 mm and >2-mm rates within 0.3 percentage points), arguing against an in-sample bias. Age was recovered in 3,774 (97.5%); after retaining one examination per child, 3,406 formed the presumed-normal cohort (median age 2.0 years; 767 infants).

Yield and selection. The 47% of examinations without a valid trace were substantially younger than the traced subset (median age 0.75 vs 2.0 years; 61% vs 21% infants; P < .001, Mann– Whitney), reflecting that the very small infant filum and conus-centered axial coverage most often defeated tracing (untraced examinations had finer in-plane voxels, 0.48 vs 0.63 mm, and similar slice thickness; Supplementary Table S1). Report-coded sign prevalence was not lower in the untraced group (tethered cord 2.6% vs 2.1%; fatty filum 0.5% vs 0.6%), arguing against preferential loss of pathology. The infant reference represents the ∼39% of infants with a traceable filum. Because untraceable fila are most plausibly the thinnest — below the resolution to segment — their omission would, if anything, bias the traced infant diameters upward; the low infant >2-mm rate is thus a conservative estimate, and the age gradient is unlikely to be an artifact of this selection.

Reference distribution. Filum diameter clustered near 1.5 mm (median 1.48 mm; 95th percentile 2.10 mm; 99th percentile 2.23 mm) and was nearly identical between sexes (median 1.48 mm in both). Age-band percentiles are given in Table 1 and Figure 1.

**Table 1.** Filum terminale diameter by age (presumed-normal cohort, n = 3,406).

| Age band | n | Median | 95th pct | 97th pct | % >2 mm<br>(95% CI)* |
| --- | --- | --- | --- | --- | --- |
| <1 y | 767 | 1.48 | 1.96 | 2.10 | 3.1 (2.1–4.6) |
| 1–3 y | 1,369 | 1.66 | 2.10 | 2.23 | 11.1 (9.6–12.9) |
| 3–6 y | 616 | 1.52 | 2.23 | 2.23 | 15.3 (12.7–18.4) |
| 6–12 y | 592 | 1.47 | 2.23 | 2.23 | 10.0 (7.8–12.7) |
| ≥12 y | 62 | 1.47 | 2.22 | 2.23 | 9.7 (4.5–19.6) |
| All | 3,406 | 1.48 | 2.10 | 2.23 | 10.0 (9.0–11.1) |

**Figure 1.**
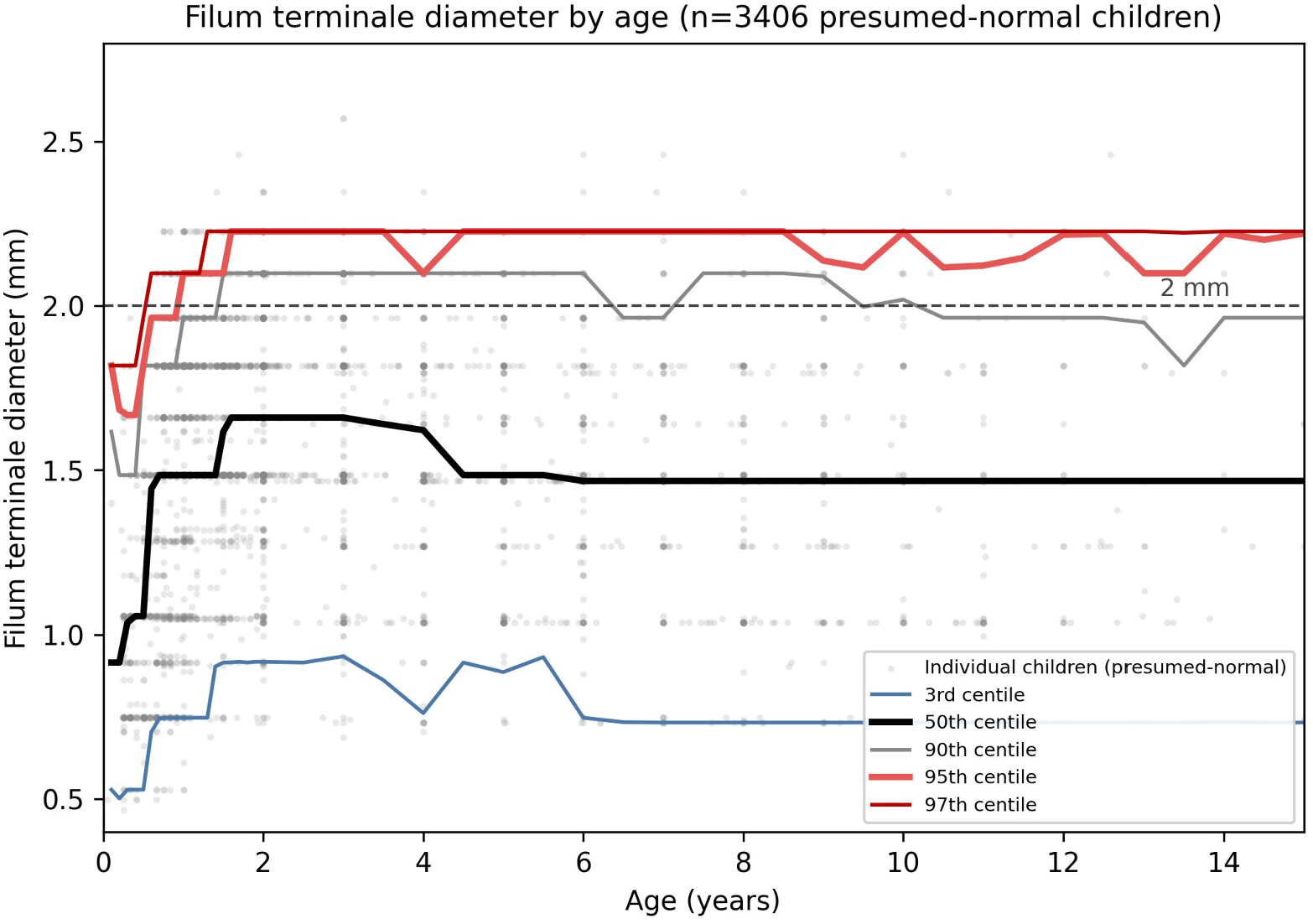
Filum terminale diameter versus age in 3,406 presumed-normal children, with empirical, discrete 3rd/50th/90th/95th/97th centiles (not a smooth biological growth curve) and the 2-mm reference line. The horizontal banding of points and the step-like centiles reflect voxel-level quantization of a 1–3-voxel structure (see Limitations); the curve is for visualization only.

The fixed 2-mm threshold is not age-stationary, and this is not a resolution artifact. In-plane voxel size correlated weakly with age (infants imaged at slightly finer resolution), so we controlled for it. Within the single dominant acquisition (in-plane voxel 0.66 mm, n = 2,757), 2 mm corresponded to the 94th percentile in infants — only 5.6% (95% CI 3.8–8.2) exceeded it — but to the 83rd percentile by 3–6 years, where 17.4% (14.4–20.8) exceeded it; the rate was 12.7% at 1–3 years and 10.6% at 6–12 years (Table 2, Figure 2). In multivariable logistic regression adjusting for both in-plane voxel size and slice thickness, the age effect was robust: compared with infants, the odds of a filum >2 mm were 2.5-fold at 1–3 years (95% CI 1.6–3.9), 4.7-fold at 3–6 years (2.8–7.7), and 3.2-fold at 6–12 years (1.8–5.6) (all P < .001); slice thickness was itself associated (odds ratio 1.22/mm, P = .003) but did not abolish the age effect. The age effect was not an artifact of the specific cutoff or of borderline quantization: it held when the threshold was set at 1.8, 2.0, or 2.2 mm (3–6 years vs infants, adjusted odds ratio 2.0, 4.7, and 7.5, respectively) and strengthened when the 1.8–2.2-mm gray zone was excluded (odds ratio 11.5; P < .001). The same fixed cutoff therefore flags roughly the top 5% of infants but one in six preschoolers, independent of acquisition resolution. We regard these resolution-controlled estimates (Table 2) as the primary result; the pooled age-band rates (Table 1) are shown for completeness and read marginally lower in infants, who were imaged at slightly finer resolution.

**Table 2.** The 2-mm threshold by age at matched image resolution.

| Age band | n (0.66 mm voxel) | % >2 mm (95% CI) | Percentile rank of 2 mm | Adjusted OR vs <1 y† |
| --- | --- | --- | --- | --- |
| <1 y | 428 | 5.6 (3.8–8.2) | 94th | 1 (ref) |
| 1–3 y | 1,187 | 12.7 (10.9–14.7) | 87th | 2.5 (1.6–3.9) |
| 3–6 y | 540 | 17.4 (14.4–20.8) | 83rd | 4.7 (2.8–7.7) |
| 6–12 y | 545 | 10.6 (8.3–13.5) | 89th | 3.2 (1.8–5.6) |
| ≥12 y | 57 | 10.5 (4.9–21.1) | 90th | 3.1 (1.1–8.5) |

Percentile rank and proportions from the single dominant acquisition (in-plane voxel 0.66 mm).
†Odds ratio for filum >2 mm from multivariable logistic regression on the full presumed-normal cohort adjusting for both in-plane voxel size and slice thickness (all $P < .05$ ; 1–3, 3–6, 6–12 years $P < .001$ ). The age effect persisted at thresholds of 1.8 and 2.2 mm and after excluding the 1.8–2.2-mm gray zone, so it is not an artifact of resolution or of the specific cutoff.

**Figure 2.**
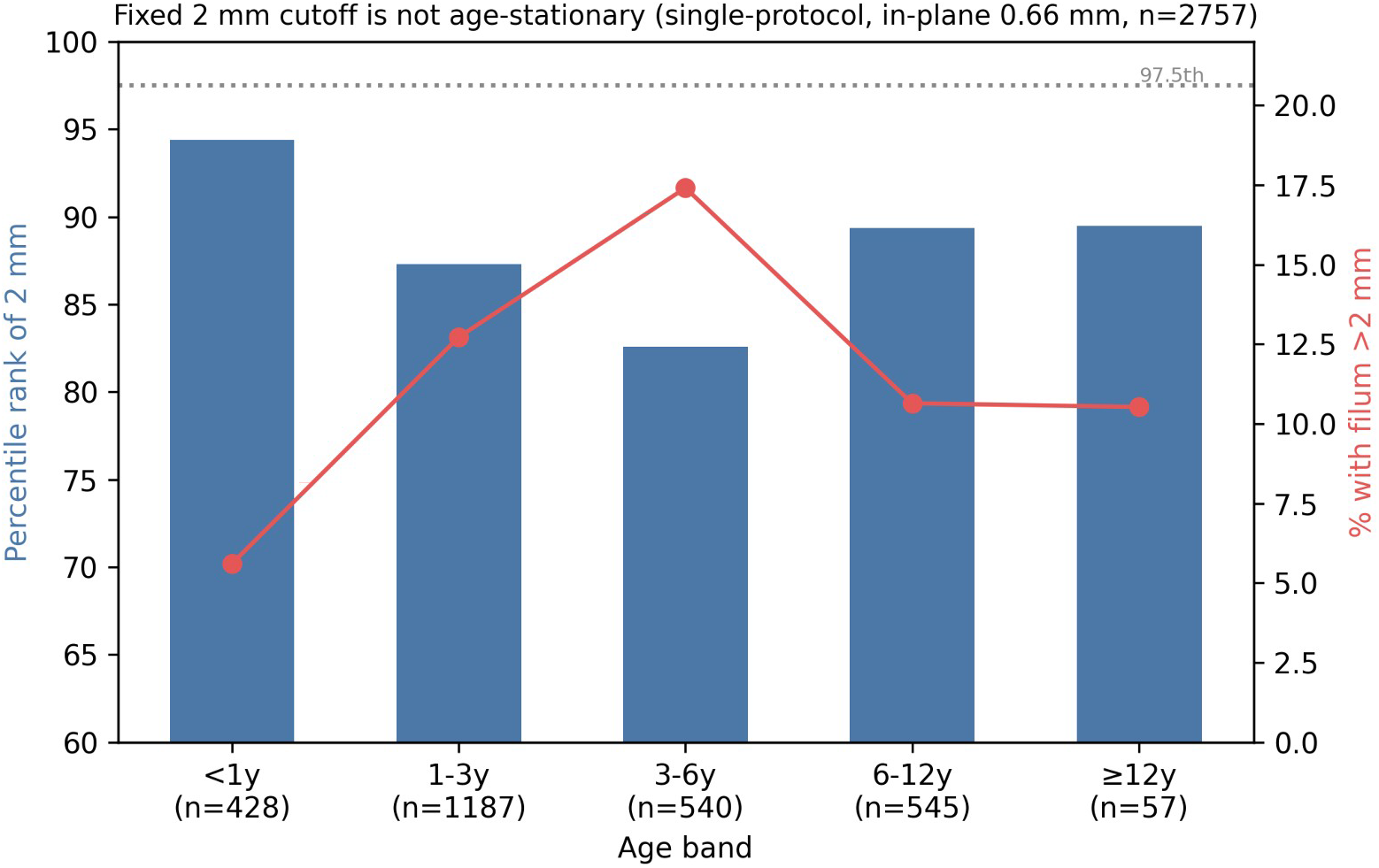
The fixed 2-mm cutoff is not age-stationary, shown at matched image resolution (single dominant acquisition, in-plane voxel 0.66 mm). Bars: percentile rank of 2 mm within each age band. Line: percentage exceeding 2 mm. The cutoff marks the ∼94th percentile in infants but the ∼83rd by 3–6 years.

Comparison groups. Report-confirmed fatty fila (n = 20) were thicker than presumed-normal fila (median 1.68 vs 1.48 mm; 35% exceeded 2 mm), confirming that the diameter measure tracks a true pathological substrate. In contrast, fila in children with a report of tethered cord (n = 61) were not thicker (median 1.30 mm), consistent with the predominantly occult presentation in this surgical population, in which the conus may be normally positioned [6], and with reports that the tethering filum is often thin and taut [8] (Figure 3). These comparison groups rest on sparse, unvalidated free-text labels and are illustrative.

**Figure 3.**
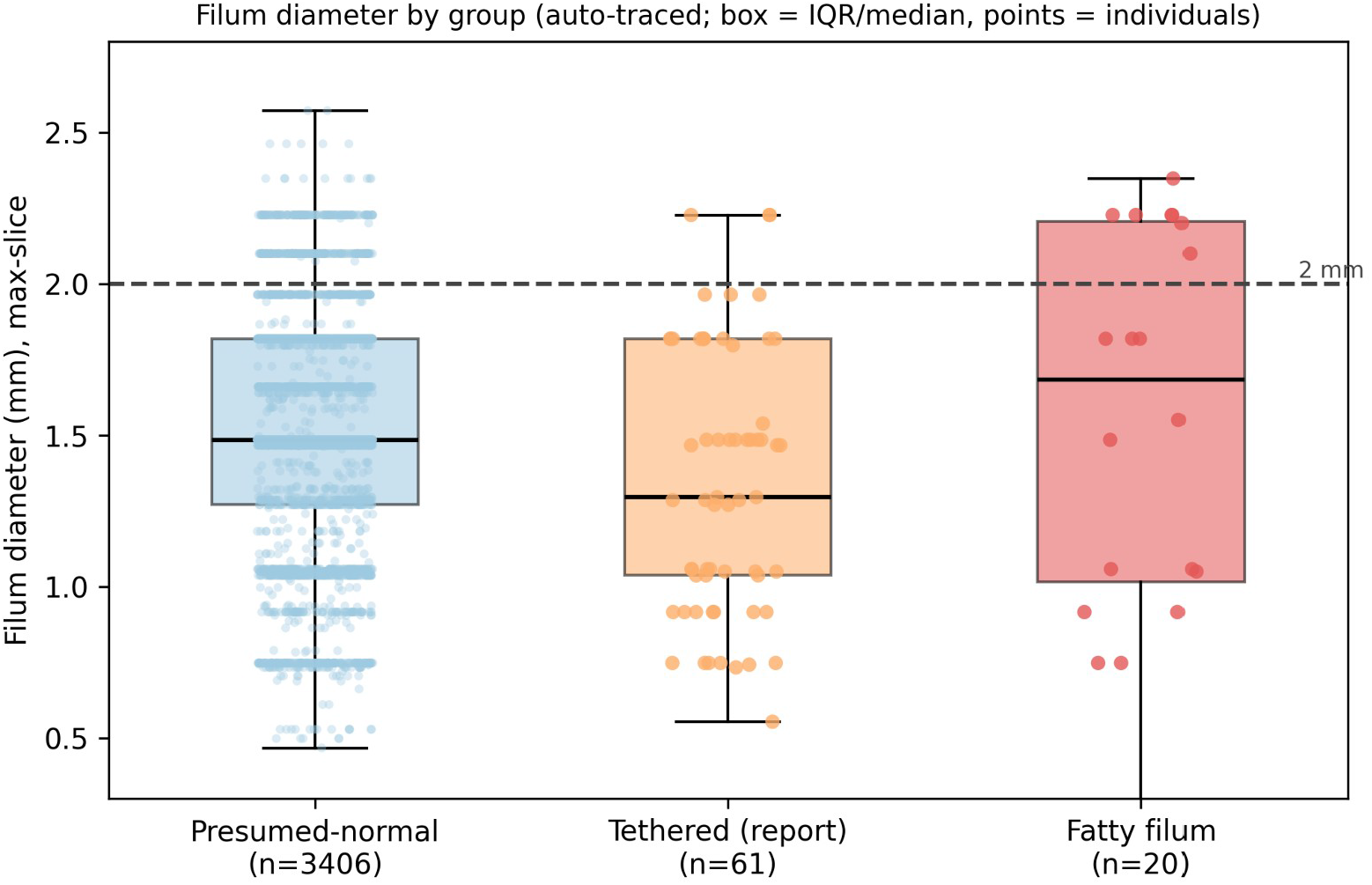
Filum diameter by group (box = median/IQR; points = individual children, shown given the small fatty group, n = 20). Report-confirmed fatty fila are shifted above the 2-mm line, whereas report-labeled tethered cord is not thicker than presumed-normal, consistent with an occult presentation.

Objective measurement versus reporting. The two rates have different denominators and are not directly equivalent: a thickened filum was recorded in the free-text report in only 3 of 9,956 examinations (0.03%, the registry-wide reporting rate), whereas a filum >2 mm was measured in ∼10% of the auto-traceable presumed-normal cohort (a resolution-dependent measurement rate; Table 2). Even allowing for the different denominators, the gap indicates that filum caliber is rarely quantified in narrative reports; objective measurement identifies a borderline-thickened group that reporting seldom captures. Because most of these fila lie just above 2 mm and the cutoff is itself age-dependent, the clinical significance of this group is uncertain.

## Discussion

In a large pediatric cohort the filum terminale diameter on routine MRI clustered near 1.5 mm — close to the adult mean [4] — but, unlike in adults [4], the 2-mm “thickened filum” threshold did not have a stationary meaning across childhood. At matched image resolution it marked roughly the top 5% of infants but ∼17% of preschoolers, and the age effect survived adjustment for in-plane voxel size and slice thickness (3–6 years vs infants, adjusted OR 4.7) and held across thresholds of 1.8–2.2 mm. A single fixed cutoff therefore behaves very differently by age — conservative in infancy, permissive in early childhood — so an age-referenced interval is a more rational basis for calling a filum thick. This parallels our companion finding for the conus, where a historically fixed anatomic criterion proved poorly aligned with the actual pediatric distribution [6].

The clinical reading of this is not that a recalibrated cutoff should drive surgery: in this population, detethering is symptom-driven; the conus can be normally positioned [6], the filum diameter frequently falls within the reference range in the present analysis, and the tethering filum can itself be thin [8]. Rather, the value is in standardized radiology reporting — flagging filum caliber against an age-appropriate reference, and specifically avoiding over-calling the borderline 2.0–2.2-mm fila that are common and usually unremarked in young children. The discordance between objective measurement (∼10% >2 mm in the traceable cohort) and free-text reporting (0.03% registry-wide) — even allowing for the different denominators — shows that caliber is rarely quantified; an objective, age-referenced measure could make reporting more consistent, while recognizing that thickening alone is neither sensitive nor specific for symptomatic tethering — filum thickness has not correlated with clinical presentation in prior series [5] — and that tethered cord remains a clinical diagnosis.

Strengths include the cohort scale, a uniform automated measurement checked against manual tracing (unbiased, though only moderately precise per filum), explicit control for the resolution confound intrinsic to a few-voxel structure (in-plane voxel, slice thickness, threshold sensitivity, gray-zone exclusion), an explicitly defined presumed-normal clinical reference, and robustness of the reference to exclusion of the tracer’s training cases.

## Limitations

First, the filum is a 1–3-voxel structure, so its diameter is quantized at approximately the in-plane voxel size and the limits of agreement against manual tracing were ±0.5 mm; absolute percentiles are correspondingly coarse and should not be over-interpreted below this resolution, and the smooth centile curve (Figure 1) is for visualization only. We mitigated, but cannot eliminate, the resolution effect by single-protocol and voxel-adjusted analyses, which preserved the age effect. Second, the automated diameter was validated on a fat-enriched paired set (n = 57); agreement specifically within the normal caliber range warrants further confirmation, ideally on higher-resolution acquisitions. Third, the tracer produced a valid trace in 53% of examinations; the untraced 47% were substantially younger (median 0.75 vs 2.0 years; 61% vs 21% infants; P < .001), as the very small infant filum and conus-centered axial coverage defeated tracing, so the infant reference represents the ∼39% of infants with a traceable filum — although, because untraceable fila are most plausibly the thinnest, this selection would bias infant diameters upward and the low infant >2-mm rate is therefore conservative. Fourth, the reference derives from clinically indicated examinations, not healthy volunteers, and is enriched for suspected spinal pathology and skewed young, so it is a large-cohort clinical reference rather than a population norm; the cohort is weighted toward infants and toddlers (median age 2.0 years), so the reference is most robust below 3 years and rests on fewer children at school age. Fifth, the comparison groups (fatty n = 20, tethered n = 61) rest on sparse free-text labels not independently validated and are illustrative only. Finally, this was a single-center, retrospective study using routine thick-slice protocols, without external validation.

## Conclusion

On routine pediatric MRI the filum terminale diameter clusters near 1.5 mm, and the fixed 2-mm “thickened filum” threshold flags roughly the top 5% of infants but ∼17% of preschoolers, even after controlling for image resolution. Filum caliber should be judged against an age-specific reference rather than a single fixed cutoff, and objective measurement identifies borderline thickening that free-text reporting rarely quantifies. A thick filum is not, by itself, a diagnosis of tethered cord, which remains a clinical determination.

## Abbreviations

TCS: tethered cord syndrome
CI: confidence interval
ICC: intraclass correlation coefficient
OR: odds ratio
nnU-Net: no-new-U-Net.

## Declarations

### Ethics approval and consent to participate

This retrospective single-center study was approved by the Ethics Committee of Women and Children’s Hospital of Ningbo University (approval no. NBFE-2026-KY-130; expedited review; approved 2 June 2026). The requirement for informed consent was waived because the study used retrospective, de-identified imaging and report data.

### Data availability

De-identified derived data (per-examination filum diameter, age band, and group label) and analysis code are available from the corresponding author on reasonable request, subject to institutional approval.

### Use of AI

Generative AI tools were used only for language editing and to assist drafting of analysis code; they were not used to generate data, results, figures, or scientific conclusions. All code outputs, statistics, and interpretations were checked and verified by the authors, who take full responsibility for the work. This disclosure is provided for transparency.

### Related manuscripts

This manuscript uses the same source registry as the conus-position preprint cited above [6]. A conus-radiomics companion manuscript has been submitted separately, and conus longitudinal reproducibility and filum T2-signal analyses are unpublished companion analyses available from the authors on request. The current manuscript has a distinct endpoint, tables, and figures.

---

**Supplementary Material — Filum Terminale Diameter on Routine Pediatric MRI: A Large-Cohort Clinical Reference in 3**,**406 Children and the Age-Dependent Meaning of the 2-mm Thickened-Filum Threshold**

**Supplementary Table S1.**
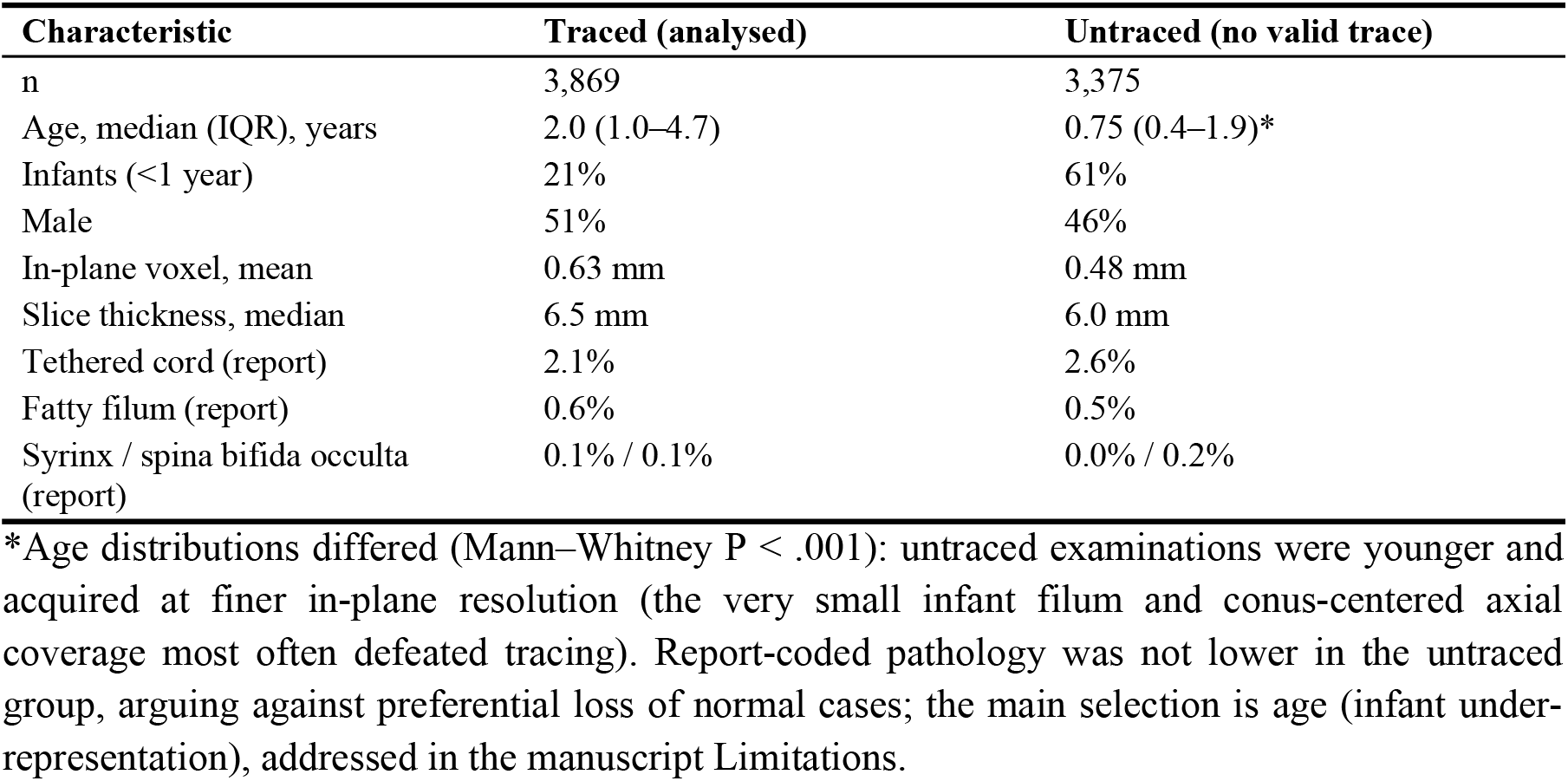
Traced versus untraced examinations.

